# Prevalence and factors associated with fear of COVID-19 in military personnel during the second epidemic wave in Peru

**DOI:** 10.1101/2023.10.03.23296474

**Authors:** Danai Valladares-Garrido, Helena Dominguez-Troncos, Cinthia Karina Picón-Reátegui, Christopher Valdiviezo-Morales, Víctor J. Vera-Ponce, Virgilio E. Failoc-Rojas, César Johan Pereira-Victorio, Darwin A. León-Figueroa, Mario J. Valladares-Garrido

**Affiliations:** Facultad de Medicina, Universidad Cesar Vallejo, Trujillo 22700, Peru; Unidad de Epidemiología y Salud Ambiental, Hospital de Apoyo II Santa Rosa, Piura 20008, Peru; School of Medicine, Universidad Nacional de Piura, Piura, Peru; Scientific Society of Medical Students from Universidad Nacional de Piura, Piura, Peru; Facultad de Medicina Humana, Universidad de San Martín de Porres, Chiclayo 15011, Peru; Instituto de Investigación en Ciencias Biomédicas, Universidad Ricardo Palma, Lima 15039, Peru; Universidad Tecnológica del Perú, Lima 15046, Peru; Research Unit for Generation and Synthesis Evidence in Health, Universidad San Ignacio de Loyola, Lima 15024, Peru; School of Medicine, Universidad Continental, Lima 15046, Peru; South American Center for Education and Research in Public Health, Universidad Norbert Wiener, Lima 15046, Peru; Oficina de Epidemiología, Hospital Regional Lambayeque, Chiclayo 14012, Peru

**Author notes:** **Correspondence:** Virgilio E. Failoc-Rojas; César Johan Pereira-Victorio.

**Keywords:** COVID-19, fear, military, Peru

## Abstract

There is few research in military members that provided protection and security during the COVID-19 crisis. We aimed to determine the prevalence and factors associated with fear of COVID-19 in military members. A cross-sectional study was conducted between November 02 and 09, 2021, during the second wave of the COVID-19 pandemic in the region of Lambayeque, Peru. The outcome was fear of COVID-19, measured with the Fear of COVID-19 Scale. The association with resilience (abbreviated CD-RISC), food insecurity (HFIAS), physical activity (IPAQ-S), eating disorder (EAT-26), and other socio-labor variables were assessed. Of 525 participants, the median age was 22, 95.8% were male, and 19.2% experienced fear of COVID-19. A higher prevalence of fear of COVID-19 was associated with age (PR=1.03; 95% CI: 1.01-1.06), religion (PP=2.05; 95% CI: 1.04-4.05), eating disorder (PR=2.95; 95% CI: 1.99-4.36), and having a relative with mental disorder (PR=2.13; 95% CI: 1.09-4.17). Overweight (PR=0.58; 95% IC: 0.37-0.90) and a high level of resilience (PR=0.63; 95% IC: 0.43-0.93) were associated with a lower prevalence of fear of COVID-19. Two out of ten military personnel were afraid of COVID-19. We recommend special attention to the factors associated with the development of suicide risk in military personnel.

## Introduction

Since its outbreak, COVID-19 has represented a threat to physical (1–4) and psychological health (5–14), which, together with its impact on the social and economic level, has had repercussions on the safety and well-being of the population (15,16). The severity (2,17) and high transmission rate of COVID-19 (18,19) has created fear in people (15,20), who, by being misinformed, have suffered the psychological impact due to the concern of becoming infected and infecting their loved ones (20,21). A total of 30.5-41.8% of fear of COVID-19 in various populations has been estimated (22,23), even a study conducted in Peruvian policemen found that 42.5% were afraid of COVID-19 (24). In Latin America, an average percentage of 15.54 on the scale of fear of COVID-19 has been described (25), which is among the lowest, compared to other continents (21.7-23.8) (23).

Fear of COVID-19 has been studied in healthcare personnel (26). However, little research has been conducted in military personnel, who have also played an important role in the fight against COVID-19 (5,27). This population has been responsible for providing protection and security to citizens, through the installation of temporary hospitals. They have ensured compliance with preventive measures and helped with the transfer of patients or corpses (28). Hence, they could be exposed to trauma and put their psychological and physical health at risk, similar to health care personnel (29). A study conducted in the Spanish Armed Forces found that 52.6% felt that they would need psychological help in the face of a new wave of the pandemic and 49.2% felt fear of death (28). Studies in the general population have found factors associated with fear of COVID-19 such as age (30), being a female (31), resilience, having chronic diseases (31,32), anxiety (33), absence of alcoholism (31), non-smoking (31), perception of getting infected with COVID-19, and fear of risk for loved ones (33). However, the instrument used to measure fear of COVID-19 was not validated (30), nor did the authors investigate other variables of interest such as religious aspects, eating disorders or family history of mental health, which we have evaluated in a military population.

Our research aims to expand the scientific evidence on fear of COVID-19 in military personnel to know the psychological impact that the pandemic has caused in this population and to allow the future implementation of psychological interventions to cope with this fear. Therefore, this study aims to determine the prevalence and factors associated with fear of COVID-19 in Peruvian military personnel during the second pandemic wave.

## Materials and Methods

### Study design and population

This study is analytical and cross-sectional. We used a secondary data analysis to identify the prevalence and the factors associated with fear of COVID-19 in military personnel in Lambayeque, Peru.

The population of the study is comprised of military personnel in charge of the first line of defense of the health emergency due to COVID-19 in the city of Lambayeque, Peru. The sample was made up of 525 military personnel. In the primary study, the military personnel that were included were those who were working for one month, at least, at the moment of the administration of the survey. In the secondary study, those who did not respond the Fear of COVID-19 Scale questions completely were excluded. This scale measured the outcome variable of this research. The sampling was non-probability, snowball-type.

### Procedure

Authorization was requested from the infantry brigade of Lambayeque to conduct the interviews with military personnel. Data collection forms were created using the REDCap system to obtain optimal data quality control and generate an online questionnaire link. The interviews were conducted in person, under strict compliance with biosecurity measures (mandatory use of masks, continuous hand sanitization, and use of open spaces to promote ventilation). Data collection was carried out from November 2 to 9, 2021, and was organized in two shifts (morning and afternoon) under the supervision of a military member of the institution. The questionnaire link was shared with the supervising military officer, who distributed it to the entire study population via text message, WhatsApp virtual message and internal coordination groups. Before starting to fill out the questionnaires, the sample members were asked for their informed consent electronically to participate in the study.

### Variable and instruments

#### Fear of COVID (Fear of COVID Scale)

The outcome variable was fear of COVID-19, operationally defined as a score higher than 16.5 in the summation of the responses obtained from the Fear of COVID-19 Scale (34). This scale consists of seven items and is reliable and valid for assessing fear of COVID-19 among the general population, with a Cronbach’s alpha of 0.82 (35). An investigation of the psychometric properties of the Spanish version of the Fear of COVID-19 Scale in a sample of the Peruvian population showed that this brief scale of fear of COVID-19 has adequate measurement properties both in terms of reliability and validity (36).

The exposure variables were the following:

#### Food Insecurity (HFIAS)

It was obtained through the Household Food Insecurity Access Scale. It is composed of nine questions that assesses food access of the last four weeks. It has adequate internal consistency (Cronbach’s alpha of 0.74) (37). Also, this scale has the following categories: food security, mild, moderate and severe food insecurity (38).

#### Physical activity (IPAQ-S)

It was obtained through the International Physical Activity Questionnaire IPAQ-S (short version). This instrument is composed of seven questions that evaluate physical activity in the last week. It is divided into low, moderate and high physical activity, after a weighted estimate of total physical activity reported in the last week (39). It shows adequate psychometric properties in Latin American population (39–41). In its abbreviated version, adequate correlations (0.26-0.69) in Spanish-speaking population have been estimated (42).

#### Eating disorder (EAT-26)

It was obtained with the EAT-26, Eating Attitudes Test, after categorizing the score obtained about the absence or presence of such disorder, using a cut-off point of 20 points. This instrument is made up of 26 items measured with a Likert scale (never, rarely, sometimes, often, very often and always). It has been validated in male population from Colombia (Cronbach’s alpha 0.98, sensitivity: 100%, specificity: 97.8%) (43)

#### Resilience (abbreviated CD-RISC)

It is operationally defined as a score above 30 points obtained from the answers of the Connor-Davidson questionnaire in the abbreviated version (44). This questionnaire is composed of ten questions and has excellent psychometric properties (Cronbach’s alpha of 0.89) in the general population (45).

#### Socio-occupational variables

Among them, we can mention age in years, gender (female, male), marital status (single, married, cohabiting, divorced), religion (none, Catholic, non-Catholic), having children (no, yes), frequent consumption of alcohol (no, yes) and tobacco (no, yes), body mass index (underweight, normal, overweight, obese), previous history of mental health (no, yes), previous family history of mental health (no, yes), report of having sought mental health support (no, yes), confidence in the government to handle the pandemic (no, yes), and how long the sample was working during the COVID-19 health emergency (one to six months, seven to twelve months, thirteen to eighteen months, nineteen months, or more).

### Statistical analysis

The statistical analysis was performed in Stata 17.

In the descriptive analysis, frequencies and percentages were estimated for the categorical variables. For age as a numerical variable, the median and interquartile range was reported, given that it had a non-normal distribution.

In the hypothesis testing analysis, we assessed whether the categorical independent variables were associated with the outcome variable (fear of COVID) using the Chi-squared test. In the case of age, we used the Mann-Whitney U test after the evaluation of the assumption of normal distribution.

We built simple and multiple regression models to investigate the factors associated with fear of COVID. Prevalence ratios (PR) and 95% confidence intervals (95% CI) were estimated. We used the generalized linear models, family Poisson distribution and log-link function with robust variance.

### Ethical aspects

The primary study was approved by the Ethics Committee of Universidad San Martín de Porres (USMP) with the code 269-2022-CIEI-FMH-USMP. The privacy of the respondents was strictly respected, and the data collected was kept anonymous. In addition, informed consent was obtained electronically from participants of legal age who were military personnel.

## Results

Regarding the 525 military personnel selected for this analysis, we found that the median age was 22, 95.8% were male, 17.1% consumed alcohol frequently, and 8.2% reported having sought mental health support due the COVID-19 pandemic. Almost half of them had food insecurity (49.5%) and 43.6% had a high level of resilience. Also, 19.2% felt fear of COVID-19. **Table 1**.

**Table 1.** Characteristics of military personnel from Lambayeque, Peru (n=525).

A total of 10.7% and 10.1% mentioned having felt fear of losing their lives due to coronavirus and having felt uncomfortable thinking about coronavirus, respectively. In addition, 9.1% reported having felt much fear of COVID-19. **Fig 1**.

**Fig 1.**
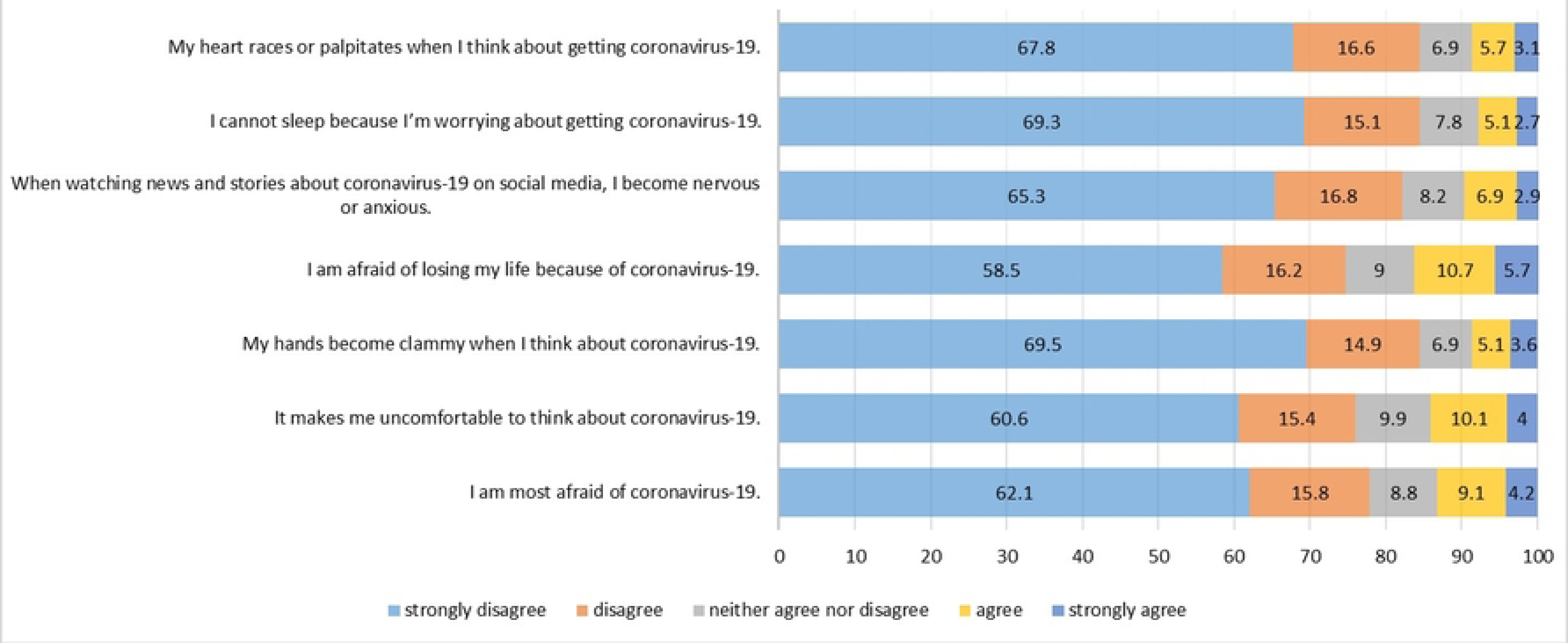
Fear of COVID-19 Scale.

We found that the participants with a low level of resilience had a higher level of prevalence of fear of COVID, in comparison with those who had a high level of resilience (23% vs. 14%; p=0.007). Moreover, military personnel with eating disorders showed a higher prevalence of fear of COVID, in comparison with those who did not have that disorder (44.4% VS. 16.4%; p< 0.001). Furthermore, age in years, religion, frequent consumption of alcohol and tobacco, having a relative with mental illnesses, and how long they have been working were associated with fear of COVID-19. **Table 2**.

**Table 2.** Factors associated with fear of COVID-19 in military personnel, bivariate analysis.

In the multiple regression model, we found that the factors associated with a high level of prevalence of fear of COVID-19 were age (in years) (PR=1.03; 95% IC: 1.01-1.06), having a religion (PP=2.05; 95% IC: 1.04-4.05), having a relative with a mental disorder (PR=2.13; 95% IC: 1.09-4.17) and having an eating disorder (PR=2.95; 95% CI: 1.99-4.36). In contrast, being overweight and having a high level of resilience reduces the prevalence of fear of COVID-19 in military personnel by 42% (PR=0.58; 95% IC: 0.37-0.90) and 37% (PR=0.63; 95% IC: 0.43-0.93). (**Table 3** and **Fig 2**).

**Fig 2.**
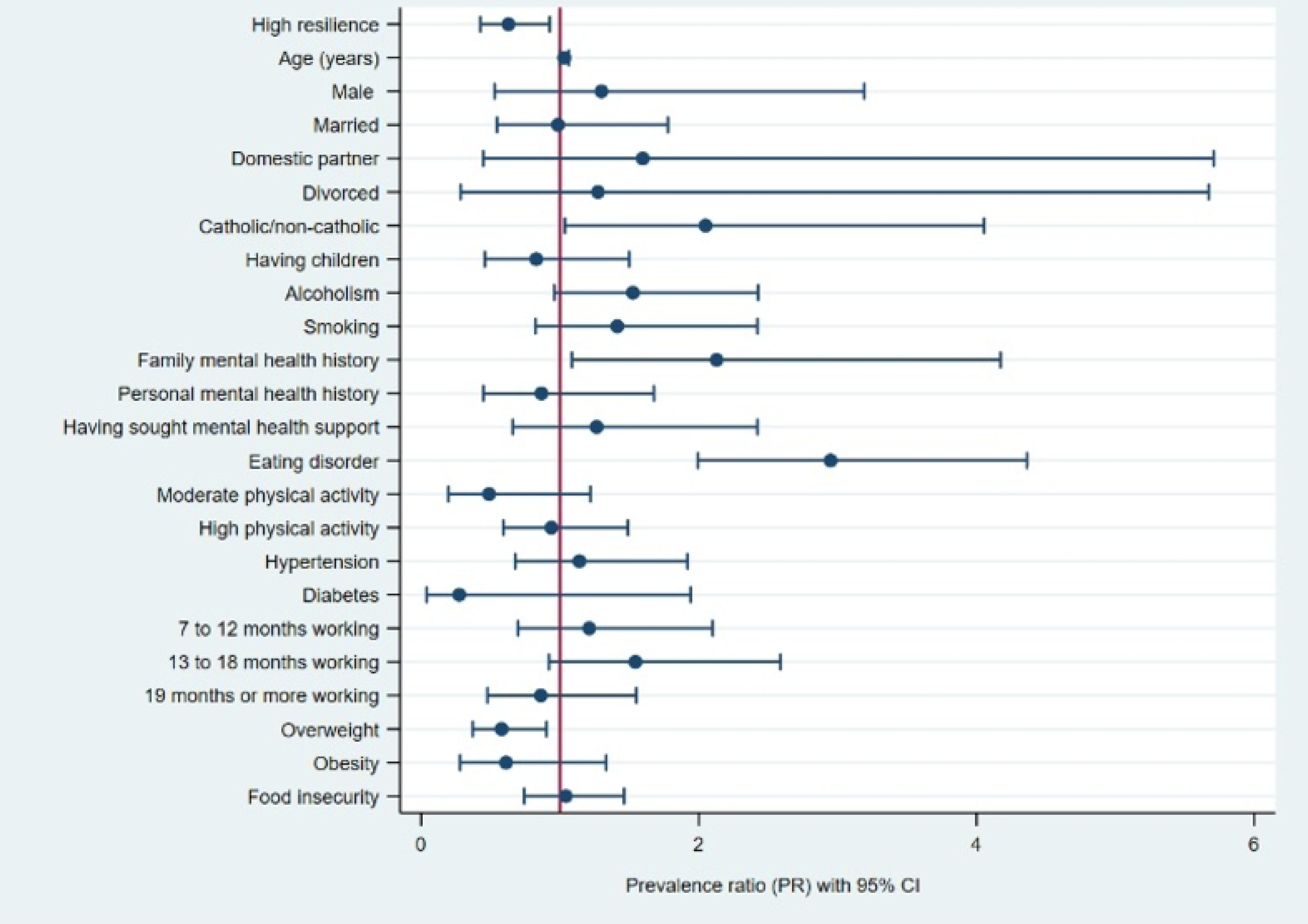
Forest plot of the factors associated with fear of COVID-19 in military personnel.

## Discussion

### Main findings

It was found that two out of ten participants were afraid of COVID-19. Factors associated with a higher prevalence of fear of COVID-19 were older age, being religious, having a family member with a history of mental illness, and having an eating disorder. Factors associated with a lower prevalence of fear of COVID-19 were being overweight and having a high level of resilience.

### Prevalence of fear of COVID-19

We found that almost two out of ten participants (19.2%) were afraid of COVID-19. This differs from what was found in our country by Caycho et al., who reported twice the incidence of fear of COVID-19 (42.5%), and 43% of the sample reported being very afraid of losing their lives to COVID-19 (46). On the other hand, in Spain, Lázaro-Perez et al. found a four times higher incidence (80%) of fear of their own or others’ death due to COVID-19 (47). The relatively low incidence of fear of COVID-19 in our study could be explained by the timing of the execution of the investigations. This is, Caycho et al. conducted their study in 2020, a time in which our country reached the maximum peaks of infected and deaths due to COVID-19; therefore, the armed and police forces played a very important role in the enforcement of security measures dictated by the Peruvian state (46). Similarly, Lázaro-Pérez et al. carried out their research at the beginning of the second wave (August-September 2020), which, together with the discovery of new variants of SARS-CoV-2 and the announcement of a new confinement, caused great consternation in the Spanish population (47).

### Resilience and fear of COVID-19

Participants with a high level of resilience had a 47% lower prevalence of fear of COVID-19. This finding coincides with what was reported by Satici et. al, who found that during the first months of the pandemic people with resilience felt less fear of COVID-19 because of a mechanism involving subjective hope and happiness in the face of adversity (48). Although we did not find a similar study that associated these variables in the population studied in the context of the COVID-19 pandemic, there is literature that supports resilience as a protective factor against mental health problems, in addition to being associated with better military performance (49,50). In addition, there is evidence suggesting that individuals with more resilience tend to externalize less fear and anxiety, which may be explained by the military training that rewards resistance, self-sufficiency and privacy, stigmatizing both physical and emotional weakness (51).

### Factors associated with fear of COVID-19

In our research, we found that the older the age, the higher the prevalence of fear of COVID-19. This is similar to what was reported in the United States by Niño et. al, who conducted a study in 10,368 American citizens. These authors identified that the older the age, the greater the perception of threat and, hence, the greater the fear of COVID-19 (52). This finding could be explained by the fact that these individuals are part of the population at risk of experiencing a more serious and complicated disease, with higher mortality rates (53–55). However, this finding differs from what was found by Soraci et al. in Italian population and Mistry SK et al. in Bangladesh, who did not find significant differences between age and fear of COVID-19 (11,12). On the contrary, Andrade et al, in Brazil, reported that the older the age, the lower the fear of COVID-19. The authors stated that the older population tends to worry less about death, has less knowledge of the disease and is generally more reluctant to accept safety measures (58).

Participants who reported being religious had a 105% higher prevalence of fear of COVID-19. This differs from the findings of Prazeres et al., in Portugal, who found no association between religiosity and fear of COVID-19 (59). On the other hand, Ghoncheh K. et al., in Iran, found that those individuals with greater religiosity and spirituality reported less fear of COVID-19 because such individuals act according to a belief system that allows them to cope with difficulties of various kinds. That helps them to decrease their levels of psychological distress. The contradictory finding of our study could be explained by work overload of the military and police forces and social distancing, which impeded the meetings of these religious communities, which deprived them of the support they obtained from their activities (masses, spiritual retreats, etc.). This last phenomenon is supported by the study conducted in the United States by Gomez et al., in which the emotional impact of the COVID-19 quarantine on people’s religiosity is analyzed (60).

It was found that overweight participants reduced prevalence of fear of COVID-19 by 42%. We did not find studies similar to ours; however, it differs from what was reported in China by Chen et al. as overweight participants showed higher levels of fear of COVID-19 compared to those with a normal BMI (61). It is based on multiple studies that relate overweight and obesity with a higher morbidity and mortality of this disease (62–64). This peculiar finding could be explained by sociodemographic and cultural factors. This is, given that, in our country and in Latin America, being overweight could erroneously not be considered a health alteration, since it would provide strength and protection against adverse situations (65,66), as in the case of the COVID-19 pandemic.

Having a family member with a mental health history increases the prevalence of fear of COVID-19 by 113%. We have not found similar studies that relate the aforementioned variables. However, this finding could be explained by evidence that states that, due to the great physical and emotional pressure triggered by their work as caregivers. Hence, family members of people with mental illnesses have a lower quality of life and are predisposed to experience anxiety, depression, post-traumatic stress disorder, among other mental problems that could justify their fear in the context of the COVID-19 pandemic (67–69).

Having an eating disorder is associated with a 195% increased prevalence of fear of COVID-19. This is similar to what was reported by Bemanian et al. in Norway and by Dos Santos et al. in Brazil, who found a strong association between psychological stress in the face of COVID-19 and social isolation with eating disorders such as emotional eating and binge eating disorder (70,71). This finding is supported by research linking such eating disorders as negative emotion management mechanisms through the release of serotonin and dopamine resulting from inadequate carbohydrate and lipid intake (72,73). On the other hand, restrictive patterns could be explained by fear of COVID-19 and concern about the quality of food and the possibility that it could be a mode of transmission (74). However, these findings differ from the studies of Ilktac et al. and Pak et al., who found a minimal association between the aforementioned variables, but when analyzed with other confounding variables in the multiple model, their effect was diluted (75,76).

### Implication of findings in mental health

Considering the great impact of the COVID-19 pandemic on mental health in the general population, the findings reported in this study support the need for the implementation of strategies focused on populations vulnerable to COVID-19, such as military personnel. Measures to address this situation could be taken through the strengthening of the Community Mental Health Centers, which should engage and establish an ongoing assessment of this population in order to avoid adverse consequences on their mental health.

### Limitations and strengths

This research has some limitations. First, its cross-sectional methodology does not allow us to attribute causality among the variables considered in the study. Second, the potential selection bias is another limitation, because the primary study included only one department, despite the fact that there is a large number of military personnel in the rest of the country’s departments. However, one of the strengths is that the probability sample obtained is ample and the instruments used have been validated in our country. In addition, variables previously not considered in similar studies such as religion, eating disorder and family history of mental problems have been considered.

### Conclusions

Our findings evidence that two out of ten military personnel felt fear of COVID-19. We recommend that special attention should be paid to the factors associated with the development of suicide risk in military personnel. Among the most important, we can mention the following: age, religion, having a family member with a history of mental illness, and having an eating disorder. Despite the limitations of our study, the results may be useful for decision-making for the creation of mental health policies and interventions at all levels of care that consider this population group as a priority in the context of the COVID-19 health emergency.

#### Ethics approval and consent to participate

The primary study was approved by the Ethics Committee of Universidad San Martín de Porres (USMP) with the code 269-2022-CIEI-FMH-USMP.

#### Consent for publication

The authors agree to allow the publication of the information contained in the submitted article.

## Availability of data and materials

The dataset generated and analyzed during the current study is not publicly available because the ethics committee has not provided permission/authorization to publicly share the data, but it is available from the corresponding author upon reasonable request.

## Author Contributions

Conceptualization, D.V.G., H.D.T., C.K.P.R., and M.J.V.G.; methodology, C.V.M., V.J.V.P., V.E.F.R., and C.J.P.V.; software, D.V.G., H.D.T., and M.J.V.G.; validation, D.V.G.; formal analysis, M.J.V.G. and D.A.L.F.; investigation, H.D.T., C.K.P.R., and M.J.V.G.; resources, D.V.G., D.A.L.F., and V.R.F.R.; data curation, M.J.V.G.; writing—original draft preparation, D.V.G., H.D.T., C.K.P.R., C.V.M., V.J.V.P., V.E.F.R., D.A.L.F., C.J.P.V and M.J.V.G.; writing—review and editing, D.V.G., H.D.T., C.K.P.R., C.V.M., V.J.V.P., V.E.F.R., D.A.L.F., C.J.P.V and M.J.V.G.; visualization, C.V.M.; supervision, M.J.V.G. All authors have read and agreed to the published version of the manuscript.

## Data Availability

The data cannot be shared publicly because the ethics committee has not given permission or authorization to publicly share the data. In addition, there are elements within the data that could allow the identification of participants. However, they are available upon reasonable request directly from the corresponding author.

## Acknowledgements

None.

## Supporting information

**S1 Table. Table 1**. Characteristics of military personnel from Lambayeque, Peru.

**S2 Table. Table 2**. Factors associated with fear of COVID-19 in military personnel, bivariate analysis.

**S3 Table. Table 3.** Factors associated with fear of COVID-19 in military personnel, simple and multiple regression analysis.

**S1 Fig. Fig 1**. Fear of COVID-19 Scale.

**S2 Fig. Fig 2**. Forest plot of the factors associated with fear of COVID-19 in military personnel.

